# Factors influencing the effectiveness of nature–based Interventions (NBIs) aimed at improving mental health and wellbeing: Protocol of an umbrella review

**DOI:** 10.1101/2022.08.05.22278412

**Authors:** Topaz Shrestha, Cheryl Voon Yi Chi, Marica Cassarino, Sarah Foley, Zelda Di Blasi

**Affiliations:** School of Applied Psychology, University College Cork, Cork, Ireland; Environmental Research Institute, University College Cork, Cork, Ireland

**Author notes:** These authors contributed equally to this work.

## Abstract

Several systematic reviews support the use of nature–based interventions (NBIs) as a mechanism of enhancing mental health and wellbeing. However, the available evidence for the effectiveness of these interventions is fragmentary and mixed. The heterogeneity of existing evidence and significant fragmentation of knowledge within the field make it difficult to draw firm conclusions regarding the effectiveness of NBIs. The aim of this mixed method umbrella review is to synthesise evidence on the effectiveness of nature–based interventions through a summative review of existing published systematic reviews and meta-analyses. A systematic search in PsycINFO, PubMed, Greenfile, Web of Science, Embase, Scopus, Academic Search Complete (EBSCO), Environment Complete (EBSCO), Cochrane Library, CINAHL, Health Policy Reference Center and Google Scholar will be performed from inception to May 2022. The search strategy will aim to find published systematic reviews of nature–based interventions (NBIs) where improving health and wellbeing is an explicit goal. This is a mixed method review and systematic reviews with both quantitative and qualitative data synthesis will be considered. Two authors will independently perform the literature search, record screening, data extraction, and quality assessment of each included systematic review and meta-analysis. The individual qualitative and quantitative syntheses will be conducted in parallel and then combined in an overarching narrative synthesis. The quantitative evidence will be used to assess the strength and direction of effect of nature–based interventions on mental health and wellbeing outcomes. Evidence drawn from qualitative studies will be analysed and synthesised to understand the various pathways to engagement, process of involvement and experiential factors which may mediate experiences. The risk of bias of the systematic reviews will be assessed using a 16-item Assessment of Multiple Systematic Reviews 2 (AMSTAR2) checklist. This review is registered on PROSPERO (CRD42022329179).

## Introduction

Connecting with nature is an important element of many people’s lives and a substantial body of research supports nature’s restorative influence on our mental health and wellbeing (1-3). More recently, the benefits of nature–based interactions are becoming increasingly acknowledged across disciplines from Positive Psychology and Urban Planning to Medicine and Public Health. This research demonstrates a consistent positive trend between engagement with nature and improved physical and mental health outcomes (4, 5). Therefore, it is of significant concern that urbanisation, environmental degradation and the challenges of modern living are leading to a reduction in engagement with the natural environment. A presiding narrative in developed nations is that modern-urbanized lifestyles have diminished healthy human relationships with natural environments resulting in a multitude of health issues and reduced wellbeing (4, 6). Many of us seem to be both physically and psychologically disconnected from nature and this has implications, for both the wellbeing of the environment and individuals (7). While long-acknowledged as practices across cultures, nature–based therapeutic interventions have grown substantially in number and type in recent years (1, 8). Western science is beginning to realise what indigenous cultures have always known – engagement with the natural environment can support, enhance and restore our health and wellbeing (5).

There is growing interdisciplinary interest in the potential for ‘nature–based interventions’ (NBIs) to assist in promoting and restoring mental health and wellbeing. Nature–based interventions can facilitate change through a relatively structured promotion of nature–based experiences. Although a generally accepted definition is lacking, NBIs can be defined as intentional programmes, activities or strategies that aim to engage people in nature–based experiences with the specific objective of enhancing health and wellbeing (9, 10). NBIs are deliberate therapeutic processes that recognize nature–human kinship (11). These interventions can be broadly categorized into those that change the environment in which people live, learn, work, recreate and heal (for example, the provision of parks in cities or gardens in hospitals) and those that alter behaviour (for example, engaging people through organized programmes such as wilderness therapy) (9). Significant variety exists in practice, from sea swimming and forest-bathing to expedition-based wilderness programmes (5, 12, 13). These interventions can be centred around green space, blue space or an amalgamation of both. Greenspace is habitually comprised of vegetation and associated with natural elements. There are two possible interpretations of greenspace. Firstly, the interpretation that greenspace refers to areas of vegetation in a landscape, such as forests and wilderness areas, gardens and backyards, street trees and parks, farmland, geological formations, coastal areas and food crops. This interpretation encompasses the overarching concept of nature, or natural areas in general. The second interpretation focuses on urban vegetation, including parks, gardens, urban forests and urban farms – usually relating to a vegetated variation of open space (12). Blue space can be defined as all visible, outdoor, natural surface waters with potential for the promotion of human health and wellbeing e.g. rivers, lakes, coasts, sea, etc. (24). Research has highlighted the specific potential for freshwater, coastal and marine ecosystems to promote and restore mental health and wellbeing (24). It is evident that there is considerable overlap between blue and green spaces, however, both green and blue spaces offer very different sensory experiences and are utilised in different ways with different health outcomes and benefits that are often overlooked and remain poorly understood. Many existing reviews of NBIs define nature exposure using metrics such as the amount of green or blue space present in any given area (e.g. number of parks with access to greenery, lakes etc.) (1, 14, 15). An inherent limitation of these metrics is that they assume exposure revolves around geographic proximity, without considering whether nearby nature was actually utilised, of good quality, or inaccessible (e.g. near a busy road crossing) (16, 17). This has resulted in a call for researchers to broaden their definition of nature exposure to also investigate different types of natural settings and their characteristics (1, 14, 18). To address this, the current review focused on NBIs where ‘nature-based’ encompasses what Bloomfield (3) refers to as “time spent outside in places defined as rich in natural beauty and/or biodiversity” (p. 82). This includes both biodiverse, unregulated, wild nature lacking human involvement (3) and publicly accessible, managed urban green spaces or blues paces (e.g. parks, gardens/allotments and man-made lakes/reservoirs) (19). Understanding what NBIs are available and the various factors influencing the effectiveness of these interventions is necessary if we are to gain a clear picture of the current state of the research.

Globally, the growing interest in the restorative potential of NBIs, within healthcare, seems to be driven by a global mental health crisis and rise of non-communicable diseases (1, 20). The issue of mental health and wellbeing is particularly relevant, with rising suicide rates and lack of funding for services highlighted internationally (21). Moreover, evidence shows that there was a significant gravitation towards natural environments during the COVID-19 pandemic and that this increased engagement with nature may have buffered the negative mental and behavioural impacts of recurrent lockdowns (22). Public health administrations are beginning to acknowledge the significance of proximity to, and engagement with, natural environments ‘as an upstream health promotion intervention for populations’ (23). The recognition of the value of nature and place as a determinant of mental health and wellbeing presents a crucial opportunity to struggling healthcare systems seeking new and cost-effective services (24).

Several comparative studies, randomized controlled trials, observational studies, and subsequently systematic reviews and meta-analyses, have been conducted to investigate the efficacy of nature–based interventions (NBIs) on mental health and wellbeing outcomes (2, 10, 25, 26). Considering that nearly 80 systematic reviews are published each day, and given the extensive number of systematic reviews assessing NBIs, it is crucial to synthesize the findings of these reviews to consolidate the evidence and better inform science and practice (9, 30, 31). Systematic reviews conducted with optimal methodological rigor can provide high-quality evidence informing further research and the development of effective policies. With the increased number of systematic reviews of NBIs available, a logical and necessary next step is to conduct an umbrella review of existing systematic reviews, allowing the findings of separate reviews to be compared and contrasted, thereby providing decision makers in healthcare with an overall synthesis of the body of information available (27, 29). This is a rapidly growing field and the recent interest in nature–based solutions (NBS) and proliferation of ‘nature–based interventions’ is surpassing the policy and knowledge base. This has resulted in a general lack of understanding regarding the practical implementation of NBIs within public planning and policy and the factors influencing the effectiveness of these interventions (9, 28). This can only limit the leveraging of natural environments to improve health and wellbeing outcomes, potentially resulting in ineffective and ill–targeted investment decisions. A higher order or meta-level synthesis is required to make sense of this evidence. This will provide a broader picture of the types of interventions available, the specific mental health and wellbeing outcomes they impact upon, the drivers and barriers to using NBIs, and the methodological quality of the existing research.

There is significant fragmentation of knowledge within the field and previous studies highlight evidence gaps concerning effectiveness of interventions (1, 9, 24). The universal application of NBIs to different groups, and the diversity of nature itself has led to significant heterogeneity of intervention designs (9). There is a need for a comprehensive overview of existing evidence regarding the effectiveness of interventions, particularly given the plurality of interventions, delivery approaches, and patient groups for which they are being used (16). Moreover, vague intervention descriptions and an absence of theoretical frameworks guiding NBI design has limited the critical appraisal of these interventions (27). A deficit of comprehensive conceptual and theoretical articulation exists specifically for nature’s contribution or role in improving wellbeing outcomes, leaving NBIs without an explicit theory of change for their application as a clinical practice. This umbrella review aims to explore the drivers influencing the effectiveness of NBIs subsequently, providing insight into the theoretical underpinnings of the nature–wellbeing relationship. The decision to include both quantitative and qualitative evidence, in this mixed method review, was based on the commitment to provide an extensive and accurate summary of the existing evidence of NBIs. It is expected that the quantitative evidence will be used to assess the strength and direction of effect of NBIs on mental health and wellbeing outcomes, thereby providing particular insight into the effectiveness of interventions. Alternatively, the qualitative studies will be used to provide a more nuanced perspective of the factors influencing the effectiveness of nature–based interventions. It is anticipated that the qualitative analysis will capture the holistic experience of nature–based interventions, for participants involved, and help to understand the experience and meaning of participation in nature–based interventions, pathways to engagement, process of involvement, and factors which may mediate their experiences. The synthesis of both quantitative and qualitative findings will provide a comprehensive overview of current evidence and will help to identify gaps in knowledge, potential quality needs and directions for future research. Further knowledge and communication about the effectiveness of interventions is likely to be a valuable precursor for their use (9).

The overall objective of this mixed method umbrella review is to synthesize the evidence on the effectiveness of nature–based interventions, aimed at enhancing mental health and wellbeing, through a summative review of existing published systematic reviews and meta-analyses. Accordingly, our specific objectives were to identify: 1) what nature–based interventions (NBIs) are available, 2) what specific mental health and wellbeing outcomes might they achieve for whom, 3) what are the drivers influencing the effectiveness of NBIs, 4) what are the barriers/limitations influencing the extent to which these interventions succeed and 5) recommendations for future research, policymaking and practice.

## Methods and analysis

### Protocol registration

The umbrella review will adhere to the predesigned protocol that has been developed based on the Preferred Reporting Items for Systematic Review and Meta-Analysis Protocols (PRISMA-P) guidelines (32) (S1 Table). This project has been registered with the International Prospective Register of Systematic Reviews (PROSPERO) (registration number CRD42022329179).

### Data sources and search strategies

We will conduct a comprehensive umbrella review of all available systematic reviews on the topic using the methodology described in previous reports (31). We will adopt the Joanna Briggs Institute (JBI) methodology for umbrella reviews, which provide further guidelines specific for synthesizing the findings from multiple reviews (29). Umbrella reviews are defined as systematic overviews of systematic reviews and/or meta-analyses, that can be used to provide a summary of the evidence from multiple research syntheses (33). The systematic overview resulting from the conduct of an umbrella review is useful to explore whether the evidence base around a topic is consistent or contradictory, and to examine the reasons for the findings (29).

A systematic search of the following twelve databases will be completed: PsycINFO, PubMed, Greenfile, Web of Science, Embase, Scopus, Academic Search Complete (EBSCO), Environment Complete (EBSCO), Cochrane Library, CINAHL, Health Policy Reference Center and Google Scholar. No date limit will be placed on the search until. The search strategy will aim to find published systematic reviews of nature–based interventions (NBIs) where improving health and wellbeing is an explicit goal. Our search strategy will be comprised of three elements. Search terms relating to (i) nature–based interventions/green or blue spaces and (ii) mental health and wellbeing outcomes will be combined with (iii) systematic review OR meta-analysis and searched for in title, abstract, and keywords. We will search databases using a set of search query including keywords and Boolean operators to retrieve the relevant literature as per the objective of this review. The search strategy consists of keywords related to the natural environment, mental health, and systematic review. The selection of search terms was based on existing theories and research defining nature–based interventions as well as initial preliminary searches for the umbrella review.

Each search term will be applied twice—initially by itself, then paired with the term “systematic review” to reduce the number of returns on some of the searches, with additional searches using hyphenated variants where appropriate. The search terms for nature–based interventions/green and blue spaces and health/wellbeing outcomes will be combined with the Boolean AND and within each group the Boolean OR will be used. Aiming for as complete coverage as possible, the search may be widened, beyond the protocol, by scanning identified articles’ bibliographies and “snowballing.” The detailed search strategy, which has been developed by the full research team in consultation with a Faculty Librarian, is available in S2 Table. The results of the search will be fully reported in the final study and presented in a flowchart following the PRISMA guidelines.

Our inclusion criteria is based on the Cochrane criteria for what constitutes a systematic review as well as the AMSTAR 2 tool for the quality assessment of systematic reviews, in order to incorporate only high quality systematic reviews (34). The AMSTAR 2 domains will be used as indicators of eligibility for our study. All systematic reviews and meta-analyses that investigate the impact of nature–based interventions (NBIs) on mental health and wellbeing will be included. The search will be limited to peer-reviewed studies published in English and results will be filtered, by study type, to include solely systematic reviews. This umbrella review will include systematic reviews, with or without meta-analysis, which review any type of nature–based intervention (NBI). Unpublished grey literature will not be included. Inclusion criteria will be restricted to studies with: defined search terms, inclusion criteria and quality assessment.

The above mentioned criteria – defined search terms, inclusion criteria, quality assessment – are fundamental components of a high quality systematic review (1). Systematic reviews which examine both randomized controlled trials (RCT) and non-randomized controlled trials and observational studies (which do not have a control group) will be included in this overview. The rationale behind this decision is that the field largely consists of non-randomized trials, and excluding systematic reviews which include non-RCT studies may result in an incomplete synthesis of findings (35). Finally, this is a mixed method review and systematic reviews with both quantitative and qualitative data synthesis will be considered.

EndNote 20 software (Thomson Reuters, Toronto, Ontario, Canada) will be used to remove duplicates and screen literature. Two researchers (TS and CVYC) will independently review titles, abstracts and full-text of eligible articles. Interrater reliability (IRR) will be reported at all three stages of screening and data extraction to ensure consistency and clarity (36). Any disagreements will be resolved by discussion or by the involvement of a third reviewer (ZDB) until consensus is reached. When titles and abstracts are insufficient to determine whether to include or exclude reviews, we will download full texts to determine eligibility. Based on the umbrella review methodology, when numerous systematic reviews provide duplicated datasets for the same comparison, the systematic review with the greatest number of studies providing study-level effect estimates will be retained for further analysis (37). The following are the detailed inclusion criteria:

### Participants

There are no age or gender restrictions for participants. Children, adolescents and adults with or without mental and/or physical health problems. The routes to participation (e.g. motivations and barriers) will be considered throughout analysis to further understand how nature–based interventions could influence health and wellbeing of participants and in what contexts. It is anticipated that the qualitative evidence will provide insight into the routes to participation in nature–based interventions.

### Interventions

In this umbrella review we will include any systematic review focused on real nature– based interventions (NBIs)/exposure to green and blue spaces. Real nature is defined as a large range of green landscapes in the indoor and outdoor environment. This includes green spaces (e.g. botanic garden, or tree canopy), indoor nature (e.g. potted plants, green walls, or flowers), or real nature views (e.g. window views) (38). For the purpose of this study, NBIs are defined as programmes, activities or strategies that aim to engage people in nature–based experiences with the specific intention of improving health and wellbeing outcomes (9).

Nature–based Interventions can be broadly categorised into (i) those that change the environment in which people live, work, learn, heal or recreate (for example, the provision of gardens in schools and hospitals or parks in cities) and (ii) those that change behaviour (for example, encouraging people to engage with nature through organized programmes or other activities, green prescriptions, forest bathing and green exercise) (9). Many existing reviews of NBIs define nature exposure using metrics such as the amount or of greenspace (1, 14, 15). An inherent limitation of these metrics is that they assume exposure revolves around geographic proximity, without considering whether nearby nature was actually utilised, of good quality, or inaccessible (e.g. near a busy road crossing) (16, 17). In order to adopt a comprehensive definition of nature exposure which considers different types of natural settings and their characteristics (1, 14, 18), the current review focused on NBIs where ‘nature-based’ encompasses what Bloomfield (3) refers to as “time spent outside in places defined as rich in natural beauty and/or biodiversity” (p. 82). This included both biodiverse, unregulated, wild nature lacking human involvement (3) and publicly accessible, managed urban greenspaces (e.g. parks and gardens/allotments) (19). All included studies must encompass NBIs which integrate explicit and purposeful nature contact, incorporating blue or green space through direct nature exposure to an authentic natural setting (e.g., walking in nature/ being in a park etc.). We will exclude interventions that examined the effects of artificial nature, virtual/simulated nature, animal therapy, animal interventions, fish tanks, or nature sounds. The justification of this revolves around our focus on ‘real nature’ (38) and our conception of NBIs where ‘nature-based’ encompasses what Bloomfield (3) refers to as “time spent outside in places defined as rich in natural beauty and/or biodiversity” (p. 82).

### Outcomes

All systematic reviews which assess the mental health and wellbeing impacts experienced by individuals following active participation in a nature–based interventions will be included. Mental health, as defined by the World Health Organization (WHO), is “a state of wellbeing in which the individual realizes his or her own abilities, can cope with the normal stresses of life, can work productively and fruitfully, and is able to make a contribution to his or her community” (39). Wellbeing encompasses positive emotions and mood, the absence of negative emotions, satisfaction with life, fulfilment, and positive functioning (40) (41). All included interventions must have the promotion of mental health and wellbeing outcomes as an explicit goal (i.e., programmes that solely aim to connect people with nature without the objective of also delivering health and wellbeing benefits will be excluded).

#### Quantitative research

Includable primary outcomes will include any recognised measure of mental health and wellbeing assessed using self-reported and objective measures. Outcomes can be defined as the psychological effects of nature–based interventions related to mental health and wellbeing (e.g. life satisfaction, quality of life, vitality, stress, anxiety, exhaustion, burnout and depression). The outcomes can be categorized as: (i) mental health indices, (ii) restoration and recovery, (iii) executive functioning/cognitive ability, (iv) work and life satisfaction, and (v) psychophysiological indicators of psychological wellbeing (e.g., cortisol levels).

#### Qualitative research

Includable qualitative study’s findings will be in the form of themes, concepts and metaphors relating to the experience, meaning and perceived impacts of nature–based interventions and any factors that help or hinder their success e.g. direct quotes, and author analysis of qualitative findings.

### Data collection and verification

We will develop a standardized form for extracting data from each systematic review. The ad hoc data extraction sheet will be developed and piloted prior to data collection and will be used to ensure a controlled analysis and data retrieval. Two authors will collect the variables listed below and cross-check the accuracy of the data. Extracted data will include:

⍰ Author(s), country of origin, year of publication.
⍰ Number of articles included
⍰ Search Terms
⍰ Type/Definition of intervention reviewed
⍰ Definition of mental health/ wellbeing outcome(s) reviewed
⍰ Quality Assessment
⍰ Quantitative findings – main findings and effect sizes
⍰ Qualitative findings – themes, concepts and metaphors relating to the experience, meaning and perceived impacts of nature–based interventions and any factors influencing effectiveness of NBIs e.g. direct quotes, and author analysis of qualitative findings

### Critical appraisal

Methodological quality of the included systematic reviews will be assessed by two independent researchers using the Assessment of Multiple Systematic Reviews 2 (AMSTAR2, an updated version of AMSTAR) tool, a 16-item checklist used to critically rate the quality of an individual systematic review as high, moderate, low and critically low based on the total score of the AMSTAR2 (34). The AMSTAR 2 tool has been updated to facilitate a more detailed assessment of systematic reviews that include both randomised and non-randomised studies of healthcare interventions. The risks of bias will be analysed in relation to the particular design, conduct, and synthesis of the systematic review. Risk of bias assessment will assess methods of randomization and intervention allocation. In the case of disagreements, a discussion will be conducted with a third reviewer to reach a consensus. In the case of insufficient or additional information, the study authors will be contacted.

### Data analysis

The strategy for data synthesis will consist of firstly extracting the quantitative and qualitative data from each review, which will be entered into the screening and data extraction table. Findings will be structured around a synthesis of the characteristics of included studies, the classification of interventions used, and the types of outcomes reported. A narrative synthesis will be used to present the potential factors influencing the effectiveness of NBIs. The umbrella review format will enable a unique form of evidence synthesis whereby the researchers can stand back and gain a comprehensive summary of the breadth of research on NBIs. The results will be reported descriptively in the text, and in tables. Where possible visual techniques will be used to present the quantitative synthesis in an accessible manner, for example for a narrative approach table (indicating factors such as study quality, strength and direction/s of results) will be used to visually represent the trends in the results. Similarly, visual techniques will be used to illustrate the nature of the qualitative data and synthesis e.g. graphs, tables, flow charts etc.

Quantitative studies will be used to appraise the strength and direction of evidence of effect. However, we anticipate a limited scope for meta-analysis due to the likelihood that many individual studies will be included in more than one review, resulting in inaccurate statistical power and a risk for misleading results. Additionally the heterogeneity of intervention type given the plurality of disciplinary origins of these interventions, delivery approaches, and patient groups for which they are being used will further impede the potential for meta-analysis. As our review considers one type of “intervention” (nature– based), however of varying composition (e.g. ecotherapy, green infrastructure, or blue environments), and its effect on several different health outcomes, we consider the challenge of dissecting each included review, extracting the results from each individual study included, and the subsequent amalgamation of the results, to be of insubstantial value given the heterogeneity in the outcome measures and the unreliable accuracy of a pooled effect estimate (42). Where the quantitative study design or outcomes are so heterogeneous as to preclude meta-analysis a narrative synthesis approach will be used (43).

Qualitative studies will be used to provide a more nuanced perspective of the factors influencing the effectiveness of nature–based interventions. It is anticipated that the qualitative analysis will capture the holistic experience of nature–based interventions, for participants involved, and help to understand the experience and meaning of participation in nature–based interventions, pathways to engagement, process of involvement and factors which may mediate their experiences. Exact methods of synthesis for the included qualitative research will depend on the nature of the evidence identified. The synthesis will be sensitive to factors which may affect the impact on wellbeing, such as the demographics of participants, the context of the activities, and the implementation and specifics of the interventions.

#### Overarching synthesis

The individual qualitative and quantitative syntheses will be conducted in parallel and then combined in an overarching narrative synthesis (43). Narrative synthesis supports the contextualised integration of diverse forms of evidence to better understand the topic of the review. This approach is particularly useful in reviews of complex intervention effectiveness such as NBIs. If data permits, the analysis will be sensitive to impacts on different groups of people (e.g. age, those with mental ill health, those recovering from specific conditions or addictions). The qualitative evidence will also be used to explore those factors which help or hinder the successful development, implementation and sustainability of the particular form of NBI for different groups of people. The combined narrative synthesis will be used to develop a conceptual model (44). The model will be grounded in formulated on the synthesised results of both the qualitative and quantitative evidence.

## Discussion

By incorporating evidence from published systematic reviews and meta-analyses, we will provide a comprehensive overview of the factors influencing effectiveness of nature-based interventions (NBIs) which are aimed at enhancing mental health and wellbeing.

The recent and rapid proliferation of ‘nature–based interventions’ (NBIs), is surpassing the policy and knowledge base. This results in challenges in understanding and evaluating their tangible impact on the publics’ mental health and wellbeing (5, 12, 24). There is a need for further evidence regarding the effectiveness of interventions, particularly given the plurality of interventions, delivery approaches, and patient groups for which they are being used. While multiple interventions exist, all proposing engagement with nature as means of enhancing mental health and wellbeing, there is a dearth of guidance as to what NBIs are available and the drivers influencing their effectiveness (9). This can only impede the leveraging of natural environments to improve mental health and wellbeing outcomes, potentially leading to ineffective and ill–targeted investment decisions. We postulate that this knowledge gap exists due to the diversity of intervention designs and therapeutic approaches. Moreover, the lack of financial prioritization allocated to cost-effective NBIs impedes the potential for such interventions to ameliorate health and wellbeing on a larger scale. With the rising prevalence of substandard mental health, and the established link between poor mental health and a myriad of other noncommunicable diseases, the general population bears a significant socioeconomic burden (45, 46). We recognise that NBIs are part of a complex system influenced by social, cultural, and political factors. Subsequently, the pathways between health and nature are unequivocally linked to health inequalities (1, 16, 47, 48). It is often the most underprivileged, i.e. people with lower socioeconomic status, that stand to benefit from access to and engagement with high quality nature (12, 16, 49). Nature–based interventions could be a cost and time-effective mechanism of enhancing wellbeing at a population level. However, a concerted and systematic effort is required to understand what factors influence the effectiveness of interventions (16). Furthermore, there is evidence that policy makers and those interested in cost-effective health improvement programmes around the world are increasingly considering supporting the promotion of ‘nature–based solutions’ (NBS) (1, 50, 51). It is therefore timely that the evidence of effectiveness of nature–based interventions is reviewed in a systematic and rigorous manner.

With the increase in the amount of systematic reviews conducted, a logical next step to provide decision makers in healthcare with the evidence they require has been the conduct of reviews of existing systematic reviews. An umbrella review was chosen to provide an overview of the evidence from multiple research syntheses through an overall examination of the body of systematic and analytic reviews. This form of evidence synthesis supports comparative analysis. This method allows us to collectively evaluate the state of the evidence in broad categories of research, which may make more sense in clinical practice rather than evaluating [them] one by one (27). The umbrella reviews’ most distinguishing feature is that only the highest level of evidence, namely other systematic reviews and meta-analyses, are considered for inclusion (29, 31). By synthesising high-level evidence of the factors influencing the effectiveness of NBIs we will gain a comprehensive overview of the strengths and weaknesses of such interventions. Thus, supporting the implementation of interventions which are more targeted and subsequently more effective. Whilst there appears to be a considerable body of literature which has sought to understand the potential mental health and wellbeing benefits of nature–based interventions, no previous umbrella review, which has addressed the factors influencing the effectiveness of interventions, was identified. Several linked reviews were identified but these were either limited in scope (e.g. focusing specifically on nature’s role in psychotherapy (4), assessing exclusively built/urban natural environments (1, 52, 53), or don’t focus explicitly on nature–based interventions e.g. exploring exposure to natural environments in general rather than intentional NBIs (2). Moreover, to the best of our knowledge, this is the first mixed method umbrella review that summarizes the factors influencing the effectiveness of nature–based interventions (NBIs) thus providing particular insight into the practical application of NBIs within public planning and policy. The focus on mental health and wellbeing outcomes in wider contexts will provide a better understanding of potential approaches and pathways which are needed to gain an evidenced-based knowledge of the benefits of NBIs.

The empirical evidence relating to our research questions, both quantitative and qualitative, will be identified, appraised and synthesized. The quantitative evidence will be used to assess the strength and direction of effect of nature–based interventions on mental health and wellbeing outcomes. Evidence drawn from qualitative studies will be used to understand the various pathways to engagement, process of involvement and factors which may mediate experiences. The aim of this umbrella review is not to repeat the searches, assessment of study eligibility, assessment of risk of bias or meta-analyses from the included reviews, but rather to provide an overall picture of findings for the specific phenomenon of NBIs. Compared with a systematic review or meta-analysis limited to one treatment comparison, an umbrella review can provide a broader picture of many treatments or intervention types (29). This is more effective to inform guidelines and clinical practice when all of the management options must be considered. This umbrella review intends to provide a resource for decision– makers, in government, non–government organisations, and other interested parties, by outlining potential interventions, the specific mental health and wellbeing outcomes they might achieve for whom, the drivers influencing the extent to which these interventions succeed, and target beneficiaries. It is expected that the findings of this review will provide a roadmap for decision–makers and support the integration of NBIs into public planning and policy.

There are some limitations inherent to our umbrella review. It is anticipated that the included systematic reviews will vary in their heterogeneity and quality. This is likely to be due to the diversity of intervention type, disciplinary origins of these interventions, delivery approaches, and patient groups for which they are being used (9, 16). In addition, heterogeneity is expected to be linked to the breadth of the aims and uses of the interventions that will be potentially includable in the review, which will range from exposure to greenspace through to specific therapeutic interventions. As a result, we anticipate a limited scope for meta-analysis. We will use the AMSTAR 2 checklist to assess the risk of bias of each included study and address the concerns around the quality of included reviews.

Despite anticipated limitations, we believe that the result of this umbrella review will benefit practitioners, landscape and urban design professionals, policy-makers and the general public. A synthesis of the evidence including the methodological quality of the research, will also be of great importance to researchers in this field.

## Data Availability

No datasets were generated or analysed during the current study. All relevant data from this study will be made available upon study completion.

## Supporting Information

**S1 Table. PRISMA-P Checklist**.

(DOC)

**S2 Table. Search Strategy**.

(DOCX)

## Authors’ contributions

**Conceptualisation:** Topaz Shrestha, Zelda Di Blasi, Sarah Foley, Marica Cassarino.

**Data curation:** Topaz Shrestha, Zelda Di Blasi, Sarah Foley, Marica Cassarino, Cheryl Voon Yi Chi.

**Formal analysis:** Topaz Shrestha, Zelda Di Blasi, Sarah Foley, Marica Cassarino.

**Investigation:** Topaz Shrestha, Zelda Di Blasi, Sarah Foley, Marica Cassarino.

**Methodology:** Topaz Shrestha, Zelda Di Blasi, Sarah Foley, Marica Cassarino and Cheryl Voon Yi Chi.

**Supervision:** Zelda Di Blasi, Sarah Foley, Marica Cassarino.

**Writing – original draft:** Topaz Shrestha.

**Writing – review & editing:** Topaz Shrestha, Zelda Di Blasi, Sarah Foley, Marica Cassarino and Cheryl Voon Yi Chi.

## References

1. Van den Bosch M, Sang ÅO. Urban natural environments as nature-based solutions for improved public health–A systematic review of reviews. Environmental research. 2017;158:373–84.

2. Hossain MM, Sultana A, Ma P, Fan Q, Sharma R, Purohit N, et al. Effects of natural environment on mental health: an umbrella review of systematic reviews and meta-analyses. 2020.

3. Bloomfield D. What makes nature-based interventions for mental health successful? BJPsych international. 2017;14(4):82–5.

4. Harper NJ, Fernee CR, Gabrielsen LE. Nature’s role in outdoor therapies: an umbrella review. International journal of environmental research and public health. 2021;18(10):5117.

5. Capaldi CA, Passmore H-A, Nisbet EK, Zelenski JM, Dopko RL. Flourishing in nature: A review of the benefits of connecting with nature and its application as a wellbeing intervention. International Journal of Wellbeing. 2015;5(4).

6. Hartig T, Mitchell R, De Vries S, Frumkin H. Nature and health. Annual review of public health. 2014;35:207–28.

7. Annerstedt van den Bosch M, Depledge MH. Healthy people with nature in mind. BMC Public Health. 2015;15(1):1–7.

8. Moeller C, King N, Burr V, Gibbs GR, Gomersall T. Nature-based interventions in institutional and organisational settings: A scoping review. International Journal of Environmental Health Research. 2018;28(3):293–305.

9. Shanahan DF, Astell–Burt T, Barber EA, Brymer E, Cox DT, Dean J, et al. Nature– based interventions for improving health and wellbeing: The purpose, the people and the outcomes. Sports. 2019;7(6):141.

10. Gritzka S, Macintyre TE, Dörfel D, Baker-Blanc JL, Calogiuri G. The effects of workplace nature-based interventions on the mental health and well-being of employees: A systematic review. Frontiers in Psychiatry. 2020;11.

11. Pretty J, Barton J. Nature-based interventions and mind–body interventions: Saving public health costs whilst increasing life satisfaction and happiness. International Journal of Environmental Research and Public Health. 2020;17(21):7769.

12. Van den Berg M, Wendel-Vos W, van Poppel M, Kemper H, van Mechelen W, Maas J. Health benefits of green spaces in the living environment: A systematic review of epidemiological studies. Urban forestry & urban greening. 2015;14(4):806–16.

13. Hunter RF, Cleland C, Cleary A, Droomers M, Wheeler BW, Sinnett D, et al. Environmental, health, wellbeing, social and equity effects of urban green space interventions: A meta-narrative evidence synthesis. Environment International. 2019;130:104923-.

14. Houlden V, Weich S, Porto de Albuquerque J, Jarvis S, Rees K. The relationship between greenspace and the mental wellbeing of adults: A systematic review. PloS one. 2018;13(9):e0203000.

15. Lachowycz K, Jones AP. Towards a better understanding of the relationship between greenspace and health: Development of a theoretical framework. Landscape and urban planning. 2013;118:62–9.

16. Wilkie S, Davinson N. Prevalence and effectiveness of nature-based interventions to impact adult health-related behaviours and outcomes: A scoping review. Landscape and Urban Planning. 2021;214:104166.

17. Holland I, DeVille NV, Browning MH, Buehler RM, Hart JE, Hipp JA, et al. Measuring nature contact: a narrative review. International Journal of Environmental Research and Public Health. 2021;18(8):4092.

18. Keniger LE, Gaston KJ, Irvine KN, Fuller RA. What are the benefits of interacting with nature? International journal of environmental research and public health. 2013;10(3):913–35.

19. Taylor L, Hochuli DF. Defining greenspace: Multiple uses across multiple disciplines. Landscape and urban planning. 2017;158:25–38.

20. Bragg R, Atkins G. A review of nature-based interventions for mental health care. Natural England Commissioned Reports. 2016;204:18.

21. Pirkis J, John A, Shin S, DelPozo-Banos M, Arya V, Analuisa-Aguilar P, et al. Suicide trends in the early months of the COVID-19 pandemic: an interrupted time-series analysis of preliminary data from 21 countries. The Lancet Psychiatry. 2021;8(7):579–88.

22. Labib SM, Browning MHEM, Rigolon A, Helbich M, James P. Nature’s contributions in coping with a pandemic in the 21st century: A narrative review of evidence during COVID-19. Science of The Total Environment. 2022;833:155095.

23. Maller C, Townsend M, Pryor A, Brown P, St Leger L. Healthy nature healthy people: ‘Contact with nature’ as an upstream health promotion intervention for populations. Health Promotion International. 2006;21(1):45–54.

24. Britton E, Kindermann G, Domegan C, Carlin C. Blue care: a systematic review of blue space interventions for health and wellbeing. Health promotion international. 2020;35(1):50–69.

25. Coventry PA, Brown JE, Pervin J, Brabyn S, Pateman R, Breedvelt J, et al. Nature-based outdoor activities for mental and physical health: Systematic review and meta-analysis. SSM - Population Health. 2021;16.

26. Djernis D, Lerstrup I, Poulsen D, Stigsdotter U, Dahlgaard J, O’Toole M. A Systematic Review and Meta-Analysis of Nature-Based Mindfulness: Effects of Moving Mindfulness Training into an Outdoor Natural Setting. International Journal of Environmental Research and Public Health. 2019;16(17).

27. Papatheodorou S. Umbrella reviews: what they are and why we need them. European journal of epidemiology. 2019;34(6):543–6.

28. van den Bogerd N, Coosje Dijkstra S, Koole SL, Seidell JC, de Vries R, Maas J. Nature in the indoor and outdoor study environment and secondary and tertiary education students’ well-being, academic outcomes, and possible mediating pathways: A systematic review with recommendations for science and practice. Health Place. 2020;66:102403.

29. Aromataris E, Fernandez R, Godfrey CM, Holly C, Khalil H, Tungpunkom P. Summarizing systematic reviews: methodological development, conduct and reporting of an umbrella review approach. JBI Evidence Implementation. 2015;13(3):132–40.

30. Hoffmann F, Allers K, Rombey T, Helbach J, Hoffmann A, Mathes T, et al. Nearly 80 systematic reviews were published each day: Observational study on trends in epidemiology and reporting over the years 2000-2019. Journal of Clinical Epidemiology. 2021;138:1–11.

31. Smith V, Devane D, Begley CM, Clarke M. Methodology in conducting a systematic review of systematic reviews of healthcare interventions. BMC medical research methodology. 2011;11(1):1–6.

32. Moher D, Shamseer L, Clarke M, Ghersi D, Liberati A, Petticrew M, et al. Preferred reporting items for systematic review and meta-analysis protocols (PRISMA-P) 2015 statement. Systematic reviews. 2015;4(1):1–9.

33. Biondi-Zoccai G. Umbrella reviews. Evidence synthesis with overviews of reviews and meta-epidemiologic studies Cham, Switzerland: Springer International. 2016.

34. Shea BJ, Reeves BC, Wells G, Thuku M, Hamel C, Moran J, et al. AMSTAR 2: a critical appraisal tool for systematic reviews that include randomised or non-randomised studies of healthcare interventions, or both. bmj. 2017;358.

35. Corazon SS, Sidenius U, Poulsen DV, Gramkow MC, Stigsdotter UK. Psycho-physiological stress recovery in outdoor nature-based interventions: A systematic review of the past eight years of research. International Journal of Environmental Research and Public Health. 2019;16(10).

36. Belur J, Tompson L, Thornton A, Simon M. Interrater reliability in systematic review methodology: exploring variation in coder decision-making. Sociological methods & research. 2021;50(2):837–65.

37. Ioannidis JP. Integration of evidence from multiple meta-analyses: a primer on umbrella reviews, treatment networks and multiple treatments meta-analyses. Cmaj. 2009;181(8):488–93.

38. van den Bogerd N, Dijkstra SC, Koole SL, Seidell JC, de Vries R, Maas J. Nature in the indoor and outdoor study environment and secondary and tertiary education students’ well-being, academic outcomes, and possible mediating pathways: A systematic review with recommendations for science and practice. Health & Place. 2020;66:102403.

39. Organization WH. Global status report on noncommunicable diseases 2014: World Health Organization; 2014.

40. Diener E. Subjective well-being: The science of happiness and a proposal for a national index. American psychologist. 2000;55(1):34.

41. La Placa V, McNaught A, Knight A. Discourse on wellbeing in research and practice. International Journal of Wellbeing. 2013;3(1).

42. Brok J, Thorlund K, Gluud C, Wetterslev J. Trial sequential analysis reveals insufficient information size and potentially false positive results in many meta-analyses. Journal of clinical epidemiology. 2008;61(8):763–9.

43. Popay J, Roberts H, Sowden A, Petticrew M, Arai L, Rodgers M, et al. Guidance on the conduct of narrative synthesis in systematic reviews. A product from the ESRC methods programme Version. 2006;1(1):b92.

44. Anderson LM, Petticrew M, Rehfuess E, Armstrong R, Ueffing E, Baker P, et al. Using logic models to capture complexity in systematic reviews. Research synthesis methods. 2011;2(1):33–42.

45. Stein DJ, Benjet C, Gureje O, Lund C, Scott KM, Poznyak V, et al. Integrating mental health with other non-communicable diseases. Bmj. 2019;364.

46. Patel V, Chatterji S. Integrating mental health in care for noncommunicable diseases: an imperative for person-centered care. Health Affairs. 2015;34(9):1498–505.

47. Kruize H, van der Vliet N, Staatsen B, Bell R, Chiabai A, Muiños G, et al. Urban green space: creating a triple win for environmental sustainability, health, and health equity through behavior change. International journal of environmental research and public health. 2019;16(22):4403.

48. Barton H, Grant M. A health map for the local human habitat. Journal of the Royal Society for the Promotion of Health. 2006;126(6):252-.

49. Twohig-Bennett C, Jones A. The health benefits of the great outdoors: A systematic review and meta-analysis of greenspace exposure and health outcomes. Environmental research. 2018;166:628–37.

50. Cohen-Shacham E, Walters G, Janzen C, Maginnis S. Nature-based solutions to address global societal challenges. IUCN: Gland, Switzerland. 2016;97:2016–36.

51. Dick J, Miller JD, Carruthers-Jones J, Dobel AJ, Carver S, Garbutt A, et al. How are nature based solutions contributing to priority societal challenges surrounding human well-being in the United Kingdom: a systematic map protocol. Environmental Evidence. 2019;8(1):1–11.

52. Bird E, Ige J, Pilkington P, Pinto A, Petrokofsky C, Burgess-Allen J. Built and natural environment planning principles for promoting health: an umbrella review. BMC public health. 2018;18(1):1–13.

53. Núñez-González S, Delgado-Ron JA, Gault C, Lara-Vinueza A, Calle-Celi D, Porreca R, et al. Overview of “systematic reviews” of the built environment’s effects on mental health. Journal of Environmental and Public Health. 2020;2020.

