## Supplemental Table 1 for "Factors influencing the effectiveness of nature–based Interventions (NBIs) aimed at improving mental health and wellbeing: Protocol of an umbrella review"

**S1 Table. PRISMA-P (Preferred Reporting Items for Systematic review and Meta-Analysis Protocols) 2015 checklist: recommended items to address in a systematic review protocol***

| Section and topic | Item No | Checklist item |
| --- | --- | --- |
| ADMINISTRATIVE INFORMATION | | |
| Title: |  |  |
| Identification | 1a | Factors influencing the effectiveness of nature–based Interventions (NBIs) aimed at improving mental health and wellbeing: Protocol of an umbrella review (Title Page) |
| Update | 1b | N/A |
| Registration | 2 | PROSPERO: CRD42022329179 (Page 8) |
| Authors: |  |  |
| Contact | 3a | All names, institutional affiliations, e-mail address of all protocol authors are provided as well as physical mailing address of corresponding author (title page) |
| Contributions | 3b | The contributions of protocol authors are listed and the guarantor of the review is identified (Page 1, 21 & 22). |
| Amendments | 4 | Amendments are not expected but all deviations will be documented and discussed (Page 15). |
| Support: |  |  |
| Sources | 5a | No specific funding or sponsorship has been provided for this review |
| Sponsor | 5b | N/A |
| Role of sponsor or funder | 5c | N/A |
| INTRODUCTION | | |
| Rationale | 6 | The rationale for the review is described in the context of what is already known and the gaps in literature (Pages 6-8). |
| Objectives | 7 | We provided our explicit objectives (Page 8) and the participants, interventions, comparators, and outcomes (PICO) on (Pages 11-13) |
| METHODS | | |
| Eligibility criteria | 8 | We explicitly described our inclusion and exclusion criteria (Page 10). |
| Information sources | 9 | We described our search strategy, databases that will be used and data sources (Pages 8-10) |
| Search strategy | 10 | We described our search strategies and databases that will be systematically explored. We also described how we will extract the data (Pages 14-16). |
| Study records: |  |  |
| Data management | 11a | We described the mechanism that will be used to manage records and data throughout the review (Pages 14-17) |
| Selection process | 11b | We clearly state the process that will be used for selecting studies (Page 10) |
| Data collection process | 11c | We described the plan of extracting data from reports (Page 14-17) |
| Data items | 12 | We listed and defined all variables for which data will be sought (Pages 13-14) |
| Outcomes and prioritization | 13 | We listed and defined all outcomes for which data will be sought, including prioritization of main and additional outcomes, with rationale (Page 13-14) |
| Risk of bias in individual studies | 14 | We described anticipated methods for assessing risk of bias of individual studies, including whether this will be done at the outcome or study level (Page 14-15) |
| Data synthesis | 15a | We described criteria under which study data will be quantitatively synthesized (Pages 15-16) |
|  | 15b | We described our plan to assess heterogeneity (Page 16) |
|  | 15c | We describe narrative synthesis and the overarching synthesis of quantitative and qualitative studies (Pages 15-17) |
|  | 15d | If quantitative synthesis is not appropriate, narrative synthesis will be used (Page 16-17) |
| Meta-bias(es) | 16 | We described the meta-bias (Page 16) |
| Confidence in cumulative evidence | 17 | We will use GRADE system as described. (Page 15) |

*** It is strongly recommended that this checklist be read in conjunction with the PRISMA-P Explanation and Elaboration (cite when available) for important clarification on the items. Amendments to a review protocol should be tracked and dated. The copyright for PRISMA-P (including checklist) is held by the PRISMA-P Group and is distributed under a Creative Commons Attribution Licence 4.0.**

*From: Shamseer L, Moher D, Clarke M, Ghersi D, Liberati A, Petticrew M, Shekelle P, Stewart L, PRISMA-P Group. Preferred reporting items for systematic review and meta-analysis protocols (PRISMA-P) 2015: elaboration and explanation. BMJ. 2015 Jan 2;349(jan02 1):g7647.*
