## Supplemental Table 2 for "Factors influencing the effectiveness of nature–based Interventions (NBIs) aimed at improving mental health and wellbeing: Protocol of an umbrella review"

**S2 Table. Search strategy and keywords for this umbrella review**

| Search query | Keywords (searched within titles, abstracts, subject headings) |
| --- | --- |
| 1) Intervention | “Nature–based Interventions” OR “NBIs” OR “Nature–based health interventions” OR “Nature interventions” OR “Nature–based activities” OR “Nature–based outdoor activities” OR “Green space” OR “Blue space” OR “Blue gym” OR “Ecotherapy” OR “Eco Therapy” OR “Eco–therapy” OR “Wilderness Therapy” OR “Nature-based therapy” OR “Nature therapy” OR “Nature-based therapeutic interventions” OR “Nature involvement” OR “Forest Bathing” OR “Shinrin-Yoku” OR “Green Care” OR OR “Nature-exposure” OR “Horticultural Therapy” OR “Wilderness Therapy” OR “Nature Play” OR “Wild Play” OR “Urban green space” OR  “natural environment” OR “natural space” OR “Restorative nature” OR “Nature contact” OR “Nature exposure”, “ “Restorative garden” OR “Healing nature” OR “Healing garden” OR “Therapeutic nature” OR “Therapeutic garden” |
| 2) Outcomes (Mental Health & Wellbeing) | “Mental health” OR “Psychological health” OR “Wellbeing” OR “Well-being” OR “Well being” OR “Happiness” OR “Life Satisfaction” OR “Quality of Life” OR “Anxiety” OR “Depression” OR “Stress” OR “Psychological stress” OR “Mental stress” OR “Vitality” OR “Burnout” OR “Exhaustion” OR “Fulfillment” |
| 3) Study Designs | Systematic review or Meta-analysis |
| 4) Combination | 1 AND 2 AND 3 |
